# The role of social network structure and function in moderate and severe social and emotional loneliness: the Dutch SaNAE study in older adults

**DOI:** 10.1101/2023.08.23.23294457

**Authors:** Lisanne CJ Steijvers, Stephanie Brinkhues, Bianca Suanet, Mandy MN Stijnen, Christian JPA Hoebe, Nicole HTM Dukers-Muijrers

**Author notes:** Corresponding author: Lisanne CJ Steijvers.

## Abstract

**Background:** Loneliness is a serious public health problem and became even more visible during the COVID-19 pandemic. Yet it is unknown which aspects of social networks are most important. Here, we evaluated social network structure and function and associations with moderate and severe social and emotional loneliness in older adults.

**Methods:** This cross-sectional study includes online questionnaire data (SaNAE cohort, August-November 2020), in independently living Dutch adults aged 40 years and older. For the separate outcomes social and emotional loneliness, associations with structural network aspects (e.g., network diversity - having various types of relationships, and density - having network members who know each other), and functional network aspects (informational, emotional, and practical social support) were assessed and risk estimates were adjusted for the number of contacts, age, educational level, level of urbanization and chronic conditions. Multivariable logistic regression analyses were stratified by sex.

**Results:** Of 3,396 participants (55% men; mean age 65 years), 18% were socially lonely which was associated with a less diverse and less dense network, living alone, feeling less connected to friends, not having a club membership, and fewer emotional supporters (men only) or informational supporters (women only). 28% were emotionally lonely, which was associated with being socially lonely, and more exclusively online (versus in-person) contacts (men only), and fewer emotional supporters (women only).

**Conclusion:** Network structure and function beyond the mere number of contacts is key in loneliness, and in particular, the types of relationships are important. Public health strategies should be sex-tailored and promote network diversity and density, club membership, informational and emotional support, and in-person contact.

## 1. Background

### 1.1. Loneliness: a public health problem

Loneliness is a serious public health problem. It leads to a higher risk of premature death and the onset and progression of a range of physical and mental health problems, such as cardiovascular diseases, infectious diseases, and depression (Holt-Lunstad et al., 2015; Steptoe et al., 2013; Valtorta et al., 2016; Wang et al., 2018). During the COVID-19 pandemic, social distancing measures to contain SARS-CoV-2 transmission have led to a reduction in daily contact and an increased risk of loneliness (Auranen et al., 2021; Feehan & Mahmud, 2021; Rumas et al., 2021). Globally, loneliness is widespread, with some countries reporting that up to one in three older people feel lonely (World Health Organization, 2022). In the Netherlands, the percentage of lonely people in the adult population is substantial, partly due to the COVID-19 pandemic (Armitage & Nellums, 2020; Van Tilburg et al., 2021). Loneliness appears to impose a heavy financial burden on society (Meisters et al., 2021).

### 1.2. Assessing loneliness

Loneliness is defined as “the unpleasant experience that occurs when a person’s network of social relationships is deficient in some important way, either quantitatively or qualitatively” (Perlman & Peplau, 1981). Weiss differentiated between social and emotional loneliness with social loneliness defined as “the absence of a broader group of contacts or an engaging social network (e.g., friends, colleagues, and people in the neighborhood)”, whereas emotional loneliness is defined as “the absence of an intimate relationship or a close emotional attachment (e.g., a partner or a best friend)” (Weiss, 1975). Instruments to assess loneliness usually do not distinguish between the two types: a validated questionnaire to assess both dimensions of loneliness is the Jong Gierveld scale (De Jong Gierveld & Van Tilburg, 2006).

### 1.3. Social networks in relation to loneliness and preventive strategies

To assess loneliness implies that we acknowledge that loneliness is related to both structure and function of a social network, in addition to other types of relevant personal, social, and structural factors. Most previous studies assessing social network aspects in relation to loneliness have emphasized social network size (a structural social network aspect), demonstrating a link between social isolation (small network size) and loneliness (Kemperman et al., 2019; Moorer & Suurmeijer, 2001). Interventions to curb loneliness will first need to identify the crucial network aspects, and use these to either prevent loneliness or alleviate its consequences. Most previous interventions have focused on promoting social interactions. Yet, merely increasing the number of social relationships that a person has, is usually not effective in social and emotional loneliness (Chater & Loewenstein, 2022). Not commonly, other structural or functional network aspects are identified. Yet, a richer description of social networks based on various structural and functional aspects can be useful in the search for effective targets for the prevention of loneliness. Social network in which an individual is embedded can be composed of many or few relationships, and of various types of relationships, supports, and modes of contact (Berkman & Glass, 2000). Most studies examining loneliness included a limited set of such social network metrics or evaluated only ‘single’ network aspects. Though recommended is to evaluate structural and functional social network aspects jointly when assessing loneliness (Deckx et al., 2015; Margelisch et al., 2017; Pronk et al., 2011), most studies also lacked such evaluation of a combination of network aspects. As a result, in depth knowledge of which social network aspects are most important in loneliness, in particular emotional and social loneliness is scarce (Dahlberg et al., 2022).

### 1.4. Structural social network aspects beyond the number of network members (network size)

Structural social network aspects include network diversity (various types of social relationships), network density (how well network members are connected), homogeneity in terms of age and sex, geographical proximity, living alone and mode of contact (Berkman & Glass, 2000). Few studies examined these other structural network aspects; family-focused (less diverse) social networks were associated with loneliness among older adults, whereas having a larger share of friends in the network was inversely associated. (Wenger, 1997; Wenger & Tucker, 2002). People with more social network members in the neighborhood are less likely to be lonely, as these network members can meet up more easily.(Cacioppo et al., 2009; Weijs-Perrée et al., 2015). Not just geographical proximity, but also who live alone are more likely to be lonely (Brittain et al., 2017; Dahlberg et al., 2022; Newall et al., 2009; Taube et al., 2013).

### 1.5. Functional social network aspects

Functional social network aspects include social support from network members, such as informational, emotional, and practical support (Berkman & Glass, 2000). Social support is important for resilience as received social support may facilitate an adequate response to a possible stressful event and thereby avoiding a physical stress response or illness (Cohen & Wills, 1985). Lack of emotional support is associated with loneliness (Dahlberg et al., 2018). Also, the type of supporting relationships (family versus non-family members) were associated with risk of emotional loneliness (Van Baarsen, 2002). Social support roles have changed during the COVID-19 pandemic as people had fewer emotional or practical supporters, but also people had to depend more on their family members or neighbors for various types of support (Ottoni et al., 2022; Steijvers et al., 2022).

### 1.6. Social connectedness

During the COVID-19 pandemic, people felt less connected with family and friends (Holaday et al., 2022). Social connectedness or the subjective feeling of being embedded within a social network is important for well-being, loneliness. Feeling less connected increases the risk for loneliness (O’Rourke et al., 2018; O’Rourke & Sidani, 2017; Santini et al., 2020).

### 1.7. The present study

The Social Network Assessment in Adults and Elderly (SaNAE) study assesses social networks in relation to health. The objective of the current study is to jointly evaluate a range of structural and functional social network aspects for their association with loneliness. Specifically the health outcomes of social or emotional loneliness were evaluated separately among older adults during the COVID-19 pandemic. These associations are examined for men and women separately, as previous studies have revealed heterogeneity of network aspects by sex (Barreto et al., 2021; Dahlberg et al., 2022; Steijvers et al., 2022). Doing so reveals possible key targets for preventive strategies in men and women for the identification of social and emotional loneliness, and to promote strategies for preventing or alleviating loneliness.

## 2. Methods

### 2.1. Ethical statement

This study was approved by the Medical Ethical Committee of the University of Maastricht (METC 2018-0698, 2019-1035, and 2020-2266). Participants gave electronic informed consent.

### 2.2. Study design and population

This cross-sectional study used data from the Dutch SaNAE cohort (www.sanae-study.nl), measured in August-November 2020 using an online questionnaire.

The SaNAE study was started in 2019 and included 5,144 participants who were independent-living Dutch adults aged 40 years or older (Steijvers et al., 2021). In August-November 2020, 5,001 participants were invited for a follow-up questionnaire of whom 67% (n=3,505) responded. Respondents were slightly older (mean difference 1.8 years, p<0.001) and differed in educational level (χ^2^ = 20.884; df = 2; *p*<0.001) compared to non-responders but did not differ in sex or network size (p>0.05). Respondents without missing data on variables of interest in this study were included in further analyses (n=3,396).

### 2.3. Moderate or severe emotional and social loneliness (outcomes in analyses)

Loneliness was assessed using the six-item De Jong Gierveld Loneliness Scale (De Jong Gierveld & Van Tilburg, 2006). The six-item scale can be used to measure unidimensional loneliness, but also to measure social and emotional loneliness. Three items for social loneliness included ‘There are plenty of people I can rely on when I have problems’, ‘There are many people whom I can trust completely’, and ‘there are enough people I feel close to’. The three items for emotional loneliness included ‘I experience a general sense of emptiness’, ‘I miss having people around’, and ‘I often feel rejected’. Answer categories included totally agree, agree, neutral, disagree, and totally disagree. The answer categories neutral, disagree, and totally disagree were counted for social loneliness while the answer categories neutral, agree, and totally agree were counted for emotional loneliness. A score of zero or one was defined as ‘not lonely’ whereas a score of two or three was defined as ‘moderately/severely lonely’. Reliability tests were performed calculating Cronbach’s alpha coefficients for social and emotional loneliness items separately.

### 2.4. Social network aspects (independent variables)

Social network aspects were measured using a name-generator questionnaire. Participants were asked to provide names of family members, friends, acquaintances, and other persons who are important to them or provide support. Additional information about network members was asked using name interpreter items. A more detailed description can be found elsewhere (Steijvers et al., 2021; Steijvers et al., 2022) and in supplementary Table 1. We include a range of structural and functional metrics (Supplementary table 1).

### 2.5. Statistical analyses

Descriptive analyses were used for sociodemographic characteristics and social network aspects of the study population. All analyses were performed for the outcome variables: social and emotional loneliness. All analyses were stratified by sex.

Various multivariable logistic regression analyses models were constructed which also included the confounding variables: age, educational level, level of urbanization and chronic conditions (Type 2 Diabetes Mellitus, asthma/COPD, and cardiovascular diseases). Models (0) were created for each social network aspect separately as independent variable adjusted for confounding variables (results not presented). Models (I) were created including all social network aspects that were statistically significant from Models (0), using stepwise backward selection and also included potentially confounding variables. Models (II) additionally included social network size. Finally, for emotional loneliness as an outcome, models (III) included a variable social loneliness (as independent determinant) since social loneliness was expected to be (in part) in the pathway between network aspects and emotional loneliness (Figure 1).

**Figure 1.**
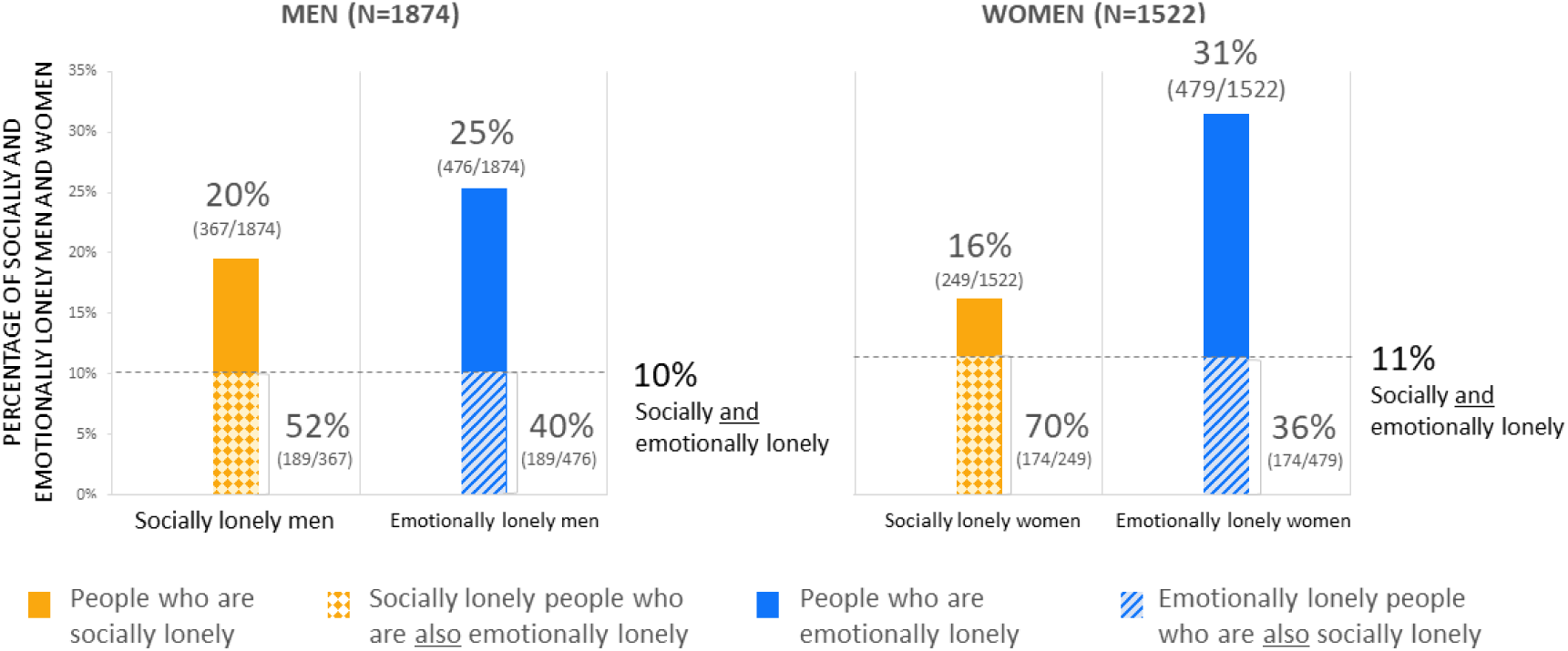
Percentages of moderate/severe social and emotional loneliness in men and women in the SaNAE study.

Before the models were built, multicollinearity between social network aspects was ruled out (correlation analyses: all correlations <0.7). A *p*-value <0.05 indicated statistical significance. All analyses were performed using IBM SPSS Statistics (version 27.0).

### 2.6. Sensitivity analyses

Sensitivity analyses were performed to assess the outcomes social and emotional loneliness when based on a different cut-off value of one and higher to reflect lonely versus not lonely (rather than the main analyses that focused on moderate and severe loneliness).

## 3. Results

### 3.1. Study population

Of all participants, 55% were men, and 43% of the participants had a theoretical educational level. The mean age was 65 years. (Table 1)

**Table 1.** Characteristics of the SaNAE study population (n=3,396)

|  | Total population | Moderate/severe social loneliness |  | Moderate/severe emotional loneliness |  |
| --- | --- | --- | --- | --- | --- |
|  | (n=3396) | Lonely (18%, n=616) | Not lonely (82%, n=2780) | Lonely (28%, n=955) | Not lonely (72%, n=2441) |
|  | % (n/N) or mean(sd) | % (n/N) or mean (sd) | % (n/N) or mean (sd) | % (n/N) or mean (sd) | % (n/N) or mean (sd) |
| **Sociodemographic characteristics** |  |  |  |  |  |
| Sex |  |  |  |  |  |
| Men | 55 (1874/3396) | 20 (367/1874) | 80 (1507/1874) | 25 (476/1874) | 75 (1398/1874) |
| Women | 45 (1522/3396) | 16 (249/1522) | 84 (1273/1522) | 32 (479/1522) | 69 (1043/1522) |
| Age (mean, sd) | 65.2 (9.8) | 65.4 (10.4) | 65.1 (9.7) | 65.8 (10.3) | 64.9 (9.7) |
| Educational level |  |  |  |  |  |
| Practical | 27 (915/3396) | 21 (192/915) | 79 (723/915) | 34 (314/915) | 66 (601/915) |
| Mixed | 30 (1009/3396) | 18 (185/1009) | 82 (824/1009) | 29 (289/1009) | 71 (720/1009) |
| Theoretical | 43 (1472/3396) | 16 (239/1472) | 84 (1233/1472) | 24 (352/1472) | 76 (1120/1472) |
| Level of urbanization |  |  |  |  |  |
| Rural | 27 (930/3396) | 17 (157/930) | 83 (773/930) | 23 (243/930) | 74 (687/930) |
| Hardly urban | 24 (810/3396) | 18 (142/810) | 82 (668/810) | 26 (213/810) | 74 (597/810) |
| Moderately urban | 19 (633/3396) | 21 (131/633) | 79 (502/633) | 29 (186/633) | 71 (447/633) |
| Strongly or extremely urban | 30 (1023/3396) | 18 (186/1023) | 82 (837/1023) | 31 (313/1023) | 69 (710/1023) |
| **Chronic conditions** |  |  |  |  |  |
| Type 2 Diabetes Mellitus (T2DM) | 9 (300) | 24 (73/300) | 76 (227/300) | 40 (121/300) | 60 (179/300) |
| Asthma/COPD | 10 (338) | 20 (68/338) | 80 (270/338) | 36 (123/338) | 64 (215/338) |
| Cardiovascular diseases | 16 (548) | 21 (114/548) | 79 (434/548) | 35 (192/548) | 65 (356/548) |
| Depression | 5 (184) | 44 (80/184) | 57 (104/184) | 72 (133/184) | 28 (51/184) |
| Score comorbidities |  |  |  |  |  |
| Zero | 68 (2313) | 16 (364/2313) | 84 (1949/2313) | 24 (544/2313) | 77 (1796/2313) |
| One | 22 (753) | 22 (169/753) | 78 (584/753) | 35 (263/753) | 65 (490/753) |
| Two or more | 10 (330) | 25 (83/330) | 75 (247/330) | 45 (148/330) | 55 (182/330) |

### 3.2. Moderate/severe social and emotional loneliness

Of the men, 20% (367/1874) were socially lonely and 25% (476/1874) were emotionally lonely. Of the women, 16% (249/1522) were socially lonely and 31% (479/1522) were emotionally lonely. Men were more likely to be socially lonely [aOR: 1.24, *p*<0.05] whereas women were more likely to be emotionally lonely [aOR: 1.50, *p*<0.001].

10% (189/1874) of the men and 11% (174/1522) of the women were both socially and emotionally lonely. Among the socially lonely men, 52% (189/367) were also emotionally lonely. For socially lonely women, this was 70% (174/249) (Figure 1).

### 3.3. Social network structure in people who are moderate or severe lonely

#### Men

Of socially lonely men, 50% had four or less network members. For 33%, their social network was composed of only family members; 20% had diverse social networks including family members, friends, and acquaintances who know each other (well) (Table 2).

**Table 2.**
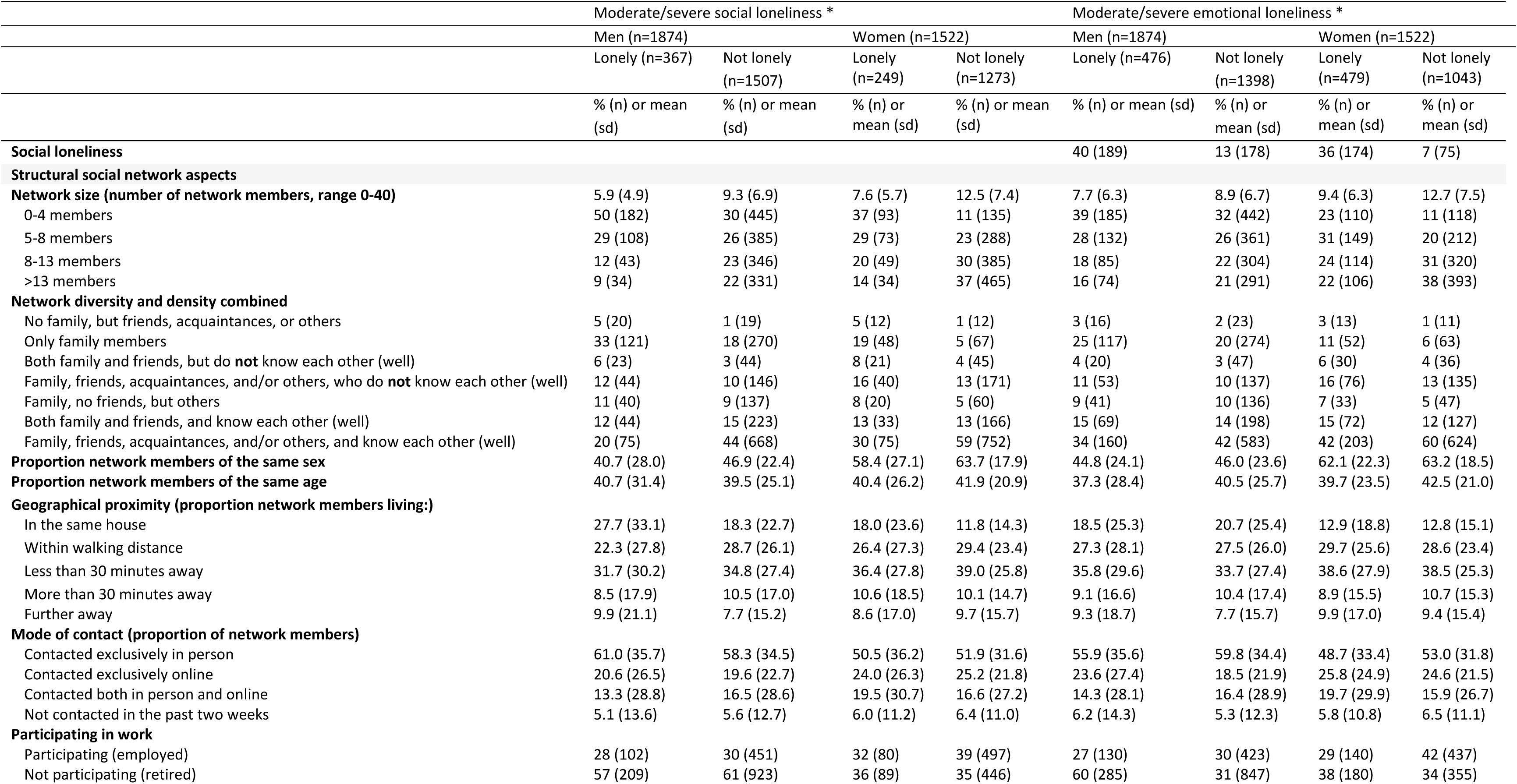
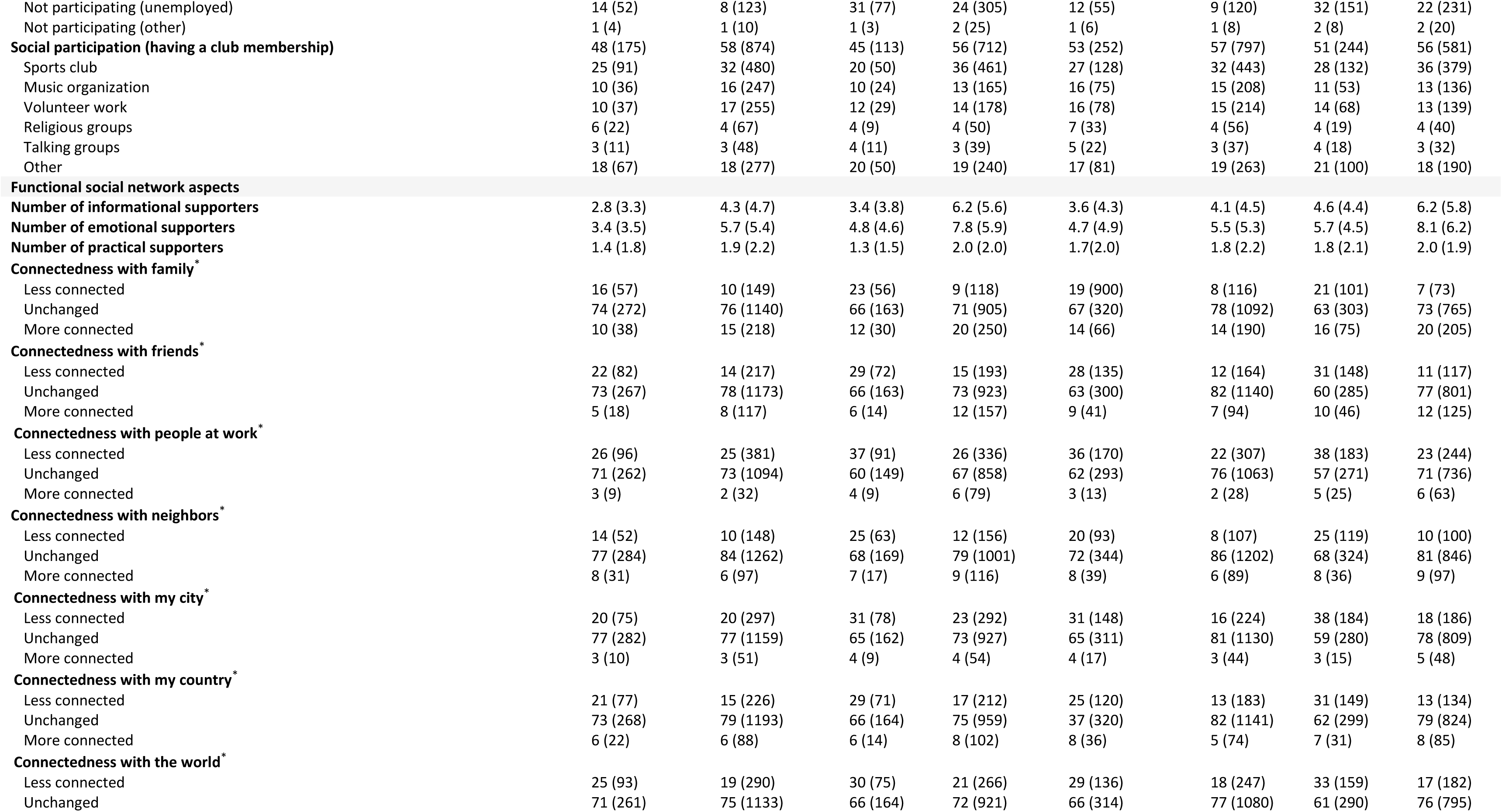
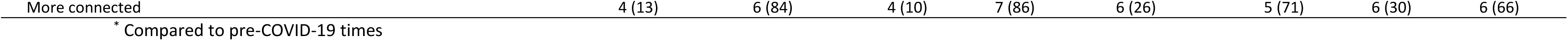
Structural and functional social network aspects and moderate/severe social and emotional loneliness of the SaNAE study population (n=3,396)

Of emotionally lonely men, 37% had four or less network members. For 19%, the social network included only family members; 30% had diverse networks.

#### Women

Of socially lonely women, 39% had four or less network members. For 25%, their social network was composed of only family members; 34% had diverse social networks including family members, friends, and acquaintances who know each other (well) (Table 2).

Of emotionally lonely women, 23% had four or less network members. For 11%, the social network included only family members; 42% had diverse networks.

### 3.4. Social network aspects associated with moderate/severe social loneliness

#### Men

In model I, independently associated with social loneliness were having less diverse and less dense social networks, a larger proportion of network members living in the same house or living far away, living alone, not participating in work, not being a member of a music organization, having fewer emotional supporters and feeling less connected with friends, and feeling more connected with neighbors (Table 3). After adding network size (model II), all these social network aspects, except social network size and not being a member of a music organization, remained associated.

**Table 3.**
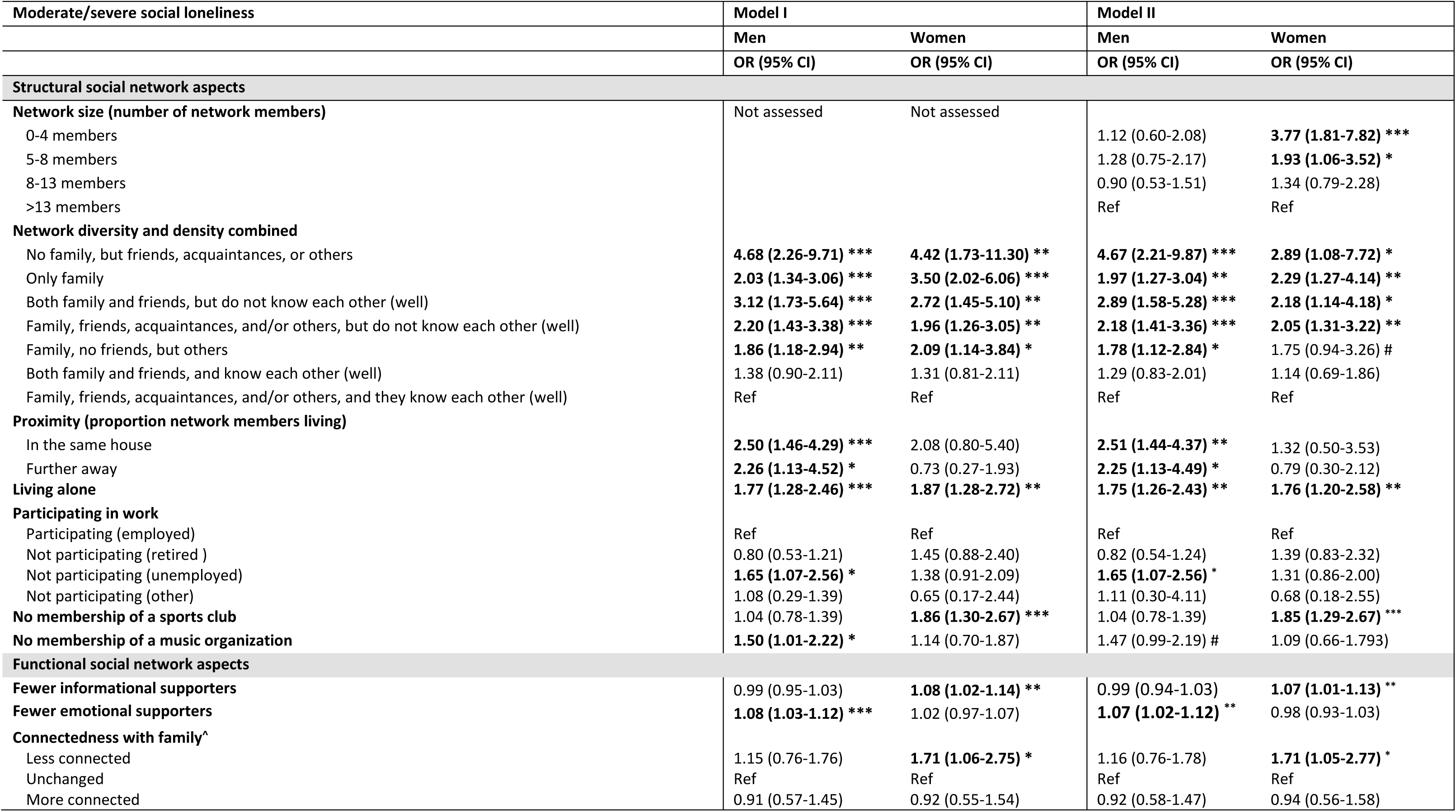
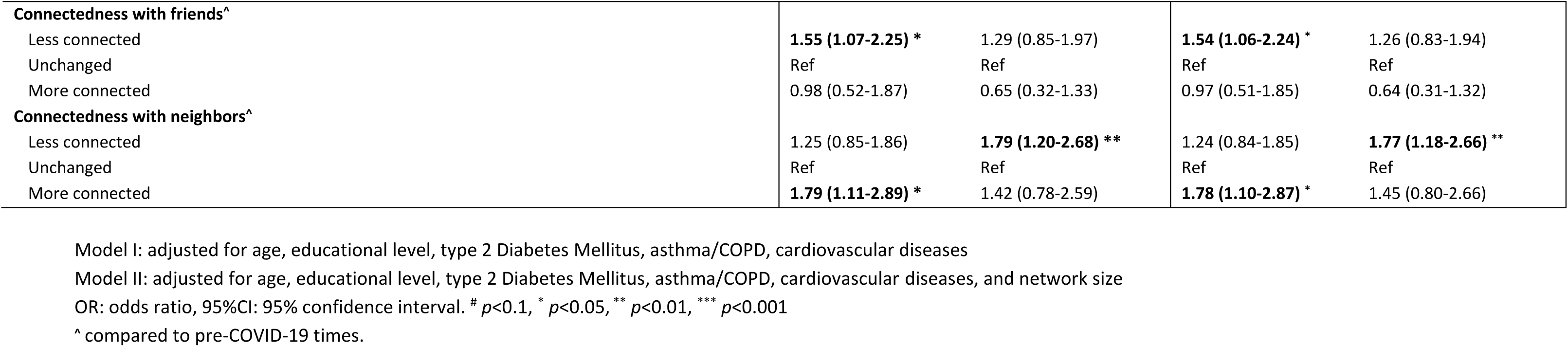
Multivariable logistic regression analyses between structural and functional social network aspects and moderate/severe social loneliness.

| Moderate/severe social loneliness | Model I |  | Model II |  |
| --- | --- | --- | --- | --- |
|  | Men | Women | Men | Women |
|  | OR (95% CI) | OR (95% CI) | OR (95% CI) | OR (95% CI) |
| **Structural social network aspects** |  |  |  |  |
| **Network size (number of network members)** | Not assessed | Not assessed |  |  |
| 0-4 members |  |  | 1.12 (0.60-2.08) | **3.77 (1.81-7.82) \*\*\*** |
| 5-8 members |  |  | 1.28 (0.75-2.17) | **1.93 (1.06-3.52) \*** |
| 8-13 members |  |  | 0.90 (0.53-1.51) | 1.34 (0.79-2.28) |
| >13 members |  |  | Ref | Ref |
| **Network diversity and density combined** |  |  |  |  |
| No family, but friends, acquaintances, or others | **4.68 (2.26-9.71) \*\*\*** | **4.42 (1.73-11.30) \*\*** | **4.67 (2.21-9.87) \*\*\*** | **2.89 (1.08-7.72) \*** |
| Only family | **2.03 (1.34-3.06) \*\*\*** | **3.50 (2.02-6.06) \*\*\*** | **1.97 (1.27-3.04) \*\*** | **2.29 (1.27-4.14) \*\*** |
| Both family and friends, but do not know each other (well) | **3.12 (1.73-5.64) \*\*\*** | **2.72 (1.45-5.10) \*\*** | **2.89 (1.58-5.28) \*\*\*** | **2.18 (1.14-4.18) \*** |
| Family, friends, acquaintances, and/or others, but do not know each other (well) | **2.20 (1.43-3.38) \*\*\*** | **1.96 (1.26-3.05) \*\*** | **2.18 (1.41-3.36) \*\*\*** | **2.05 (1.31-3.22) \*\*** |
| Family, no friends, but others | **1.86 (1.18-2.94) \*\*** | **2.09 (1.14-3.84) \*** | **1.78 (1.12-2.84) \*** | 1.75 (0.94-3.26) # |
| Both family and friends, and know each other (well) | 1.38 (0.90-2.11) | 1.31 (0.81-2.11) | 1.29 (0.83-2.01) | 1.14 (0.69-1.86) |
| Family, friends, acquaintances, and/or others, and they know each other (well) | Ref | Ref | Ref | Ref |
| **Proximity (proportion network members living)** |  |  |  |  |
| In the same house | **2.50 (1.46-4.29) \*\*\*** | 2.08 (0.80-5.40) | **2.51 (1.44-4.37) \*\*** | 1.32 (0.50-3.53) |
| Further away | **2.26 (1.13-4.52) \*** | 0.73 (0.27-1.93) | **2.25 (1.13-4.49) \*** | 0.79 (0.30-2.12) |
| **Living alone** | **1.77 (1.28-2.46) \*\*\*** | **1.87 (1.28-2.72) \*\*** | **1.75 (1.26-2.43) \*\*** | **1.76 (1.20-2.58) \*\*** |
| **Participating in work** |  |  |  |  |
| Participating (employed) | Ref | Ref | Ref | Ref |
| Not participating (retired ) | 0.80 (0.53-1.21) | 1.45 (0.88-2.40) | 0.82 (0.54-1.24) | 1.39 (0.83-2.32) |
| Not participating (unemployed) | **1.65 (1.07-2.56) \*** | 1.38 (0.91-2.09) | **1.65 (1.07-2.56) \*** | 1.31 (0.86-2.00) |
| Not participating (other) | 1.08 (0.29-1.39) | 0.65 (0.17-2.44) | 1.11 (0.30-4.11) | 0.68 (0.18-2.55) |
| **No membership of a sports club** | 1.04 (0.78-1.39) | **1.86 (1.30-2.67) \*\*\*** | 1.04 (0.78-1.39) | **1.85 (1.29-2.67) \*\*\*** |
| **No membership of a music organization** | **1.50 (1.01-2.22) \*** | 1.14 (0.70-1.87) | 1.47 (0.99-2.19) # | 1.09 (0.66-1.793) |
| **Functional social network aspects** |  |  |  |  |
| **Fewer informational supporters** | 0.99 (0.95-1.03) | **1.08 (1.02-1.14) \*\*** | 0.99 (0.94-1.03) | **1.07 (1.01-1.13) \*\*** |
| **Fewer emotional supporters** | **1.08 (1.03-1.12) \*\*\*** | 1.02 (0.97-1.07) | **1.07 (1.02-1.12) \*\*** | 0.98 (0.93-1.03) |
| **Connectedness with family^** |  |  |  |  |
| Less connected | 1.15 (0.76-1.76) | **1.71 (1.06-2.75) \*** | 1.16 (0.76-1.78) | **1.71 (1.05-2.77) \*** |
| Unchanged | Ref | Ref | Ref | Ref |
| More connected | 0.91 (0.57-1.45) | 0.92 (0.55-1.54) | 0.92 (0.58-1.47) | 0.94 (0.56-1.58) |
| **Connectedness with friends<sup>^</sup>** |  |  |  |  |
| Less connected | **1.55 (1.07-2.25) \*** | 1.29 (0.85-1.97) | **1.54 (1.06-2.24) \*** | 1.26 (0.83-1.94) |
| Unchanged | Ref | Ref | Ref | Ref |
| More connected | 0.98 (0.52-1.87) | 0.65 (0.32-1.33) | 0.97 (0.51-1.85) | 0.64 (0.31-1.32) |
| **Connectedness with neighbors<sup>^</sup>** |  |  |  |  |
| Less connected | 1.25 (0.85-1.86) | **1.79 (1.20-2.68) \*\*** | 1.24 (0.84-1.85) | **1.77 (1.18-2.66) \*\*** |
| Unchanged | Ref | Ref | Ref | Ref |
| More connected | **1.79 (1.11-2.89) \*** | 1.42 (0.78-2.59) | **1.78 (1.10-2.87) \*** | 1.45 (0.80-2.66) |

Model I: adjusted for age, educational level, type 2 Diabetes Mellitus, asthma/COPD, cardiovascular diseases
Model II: adjusted for age, educational level, type 2 Diabetes Mellitus, asthma/COPD, cardiovascular diseases, and network size
OR: odds ratio, 95%CI: 95% confidence interval. # $p < 0.1$ , \* $p < 0.05$ , \*\* $p < 0.01$ , \*\*\* $p < 0.001$
<sup>^</sup> compared to pre-COVID-19 times.

#### Women

In model I, independently associated with social loneliness were having less diverse and less dense social networks, living alone, not being a member of a sports club, having fewer informational supporters, and feeling less connected with family and neighbors. After adding network size (model II), all these social network aspects remained associated. Smaller social network size was also associated with social loneliness. (Table 3)

### 3.5. Social network aspects associated with moderate/severe emotional loneliness

#### Men

In model I, independently associated with emotional loneliness were having less diverse and less dense social networks, contacting a larger proportion of network members exclusively online, living alone, membership of a religious group, feeling less connected to friends, people from work and the city, and feeling more connected with friends, and the country. After adding network size (model II), all these social network aspects remained associated, except for network diversity and density. After adding social loneliness (model III), all social network aspects remained associated, except feeling more connected to the country. Social loneliness was also associated with emotional loneliness. (Table 4)

**Table 4.**
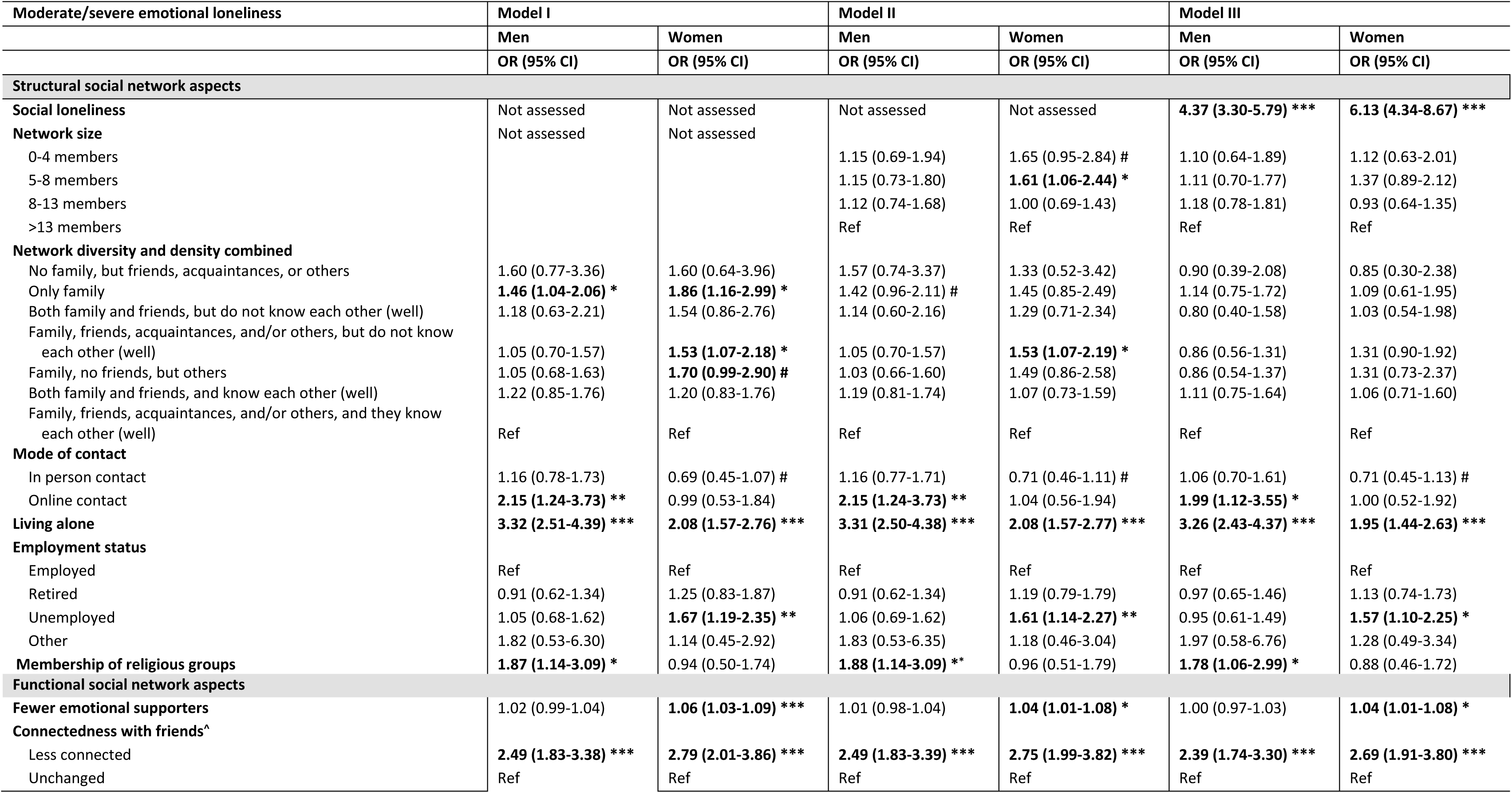
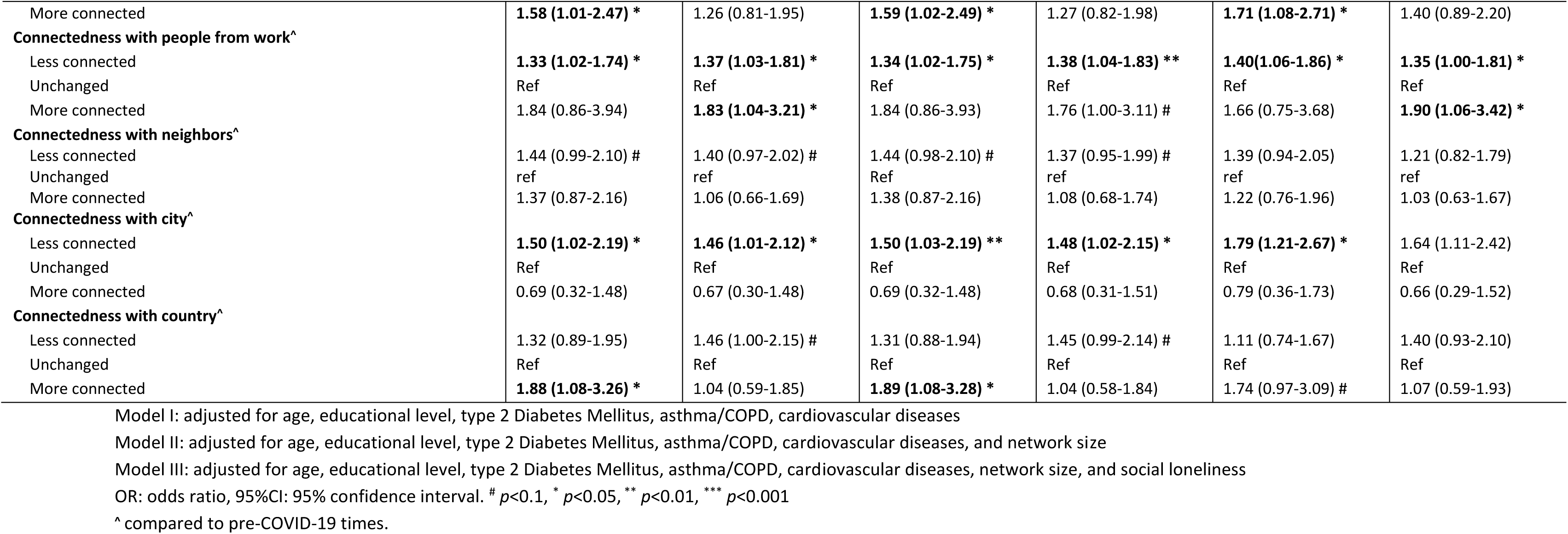
Multivariable logistic regression analyses between structural and functional social network aspects and moderate/severe emotional loneliness.

#### Women

In model I, independently associated with emotional loneliness were less diverse and less dense social networks, living alone, not participating in work, having fewer emotional supporters, and feeling less connected with friends, people from work, and the city, and feeling more connected with people from work. After adding network size (model II), all these social network aspects remained associated except feeling more connected to people from work. Smaller network size was also associated. After adding social loneliness (model III), all social network aspects remained associated except network size and diversity and density.

An visual overview of all associations between social networks and social and emotional loneliness is available in Figure 2.

**Figure 2.**
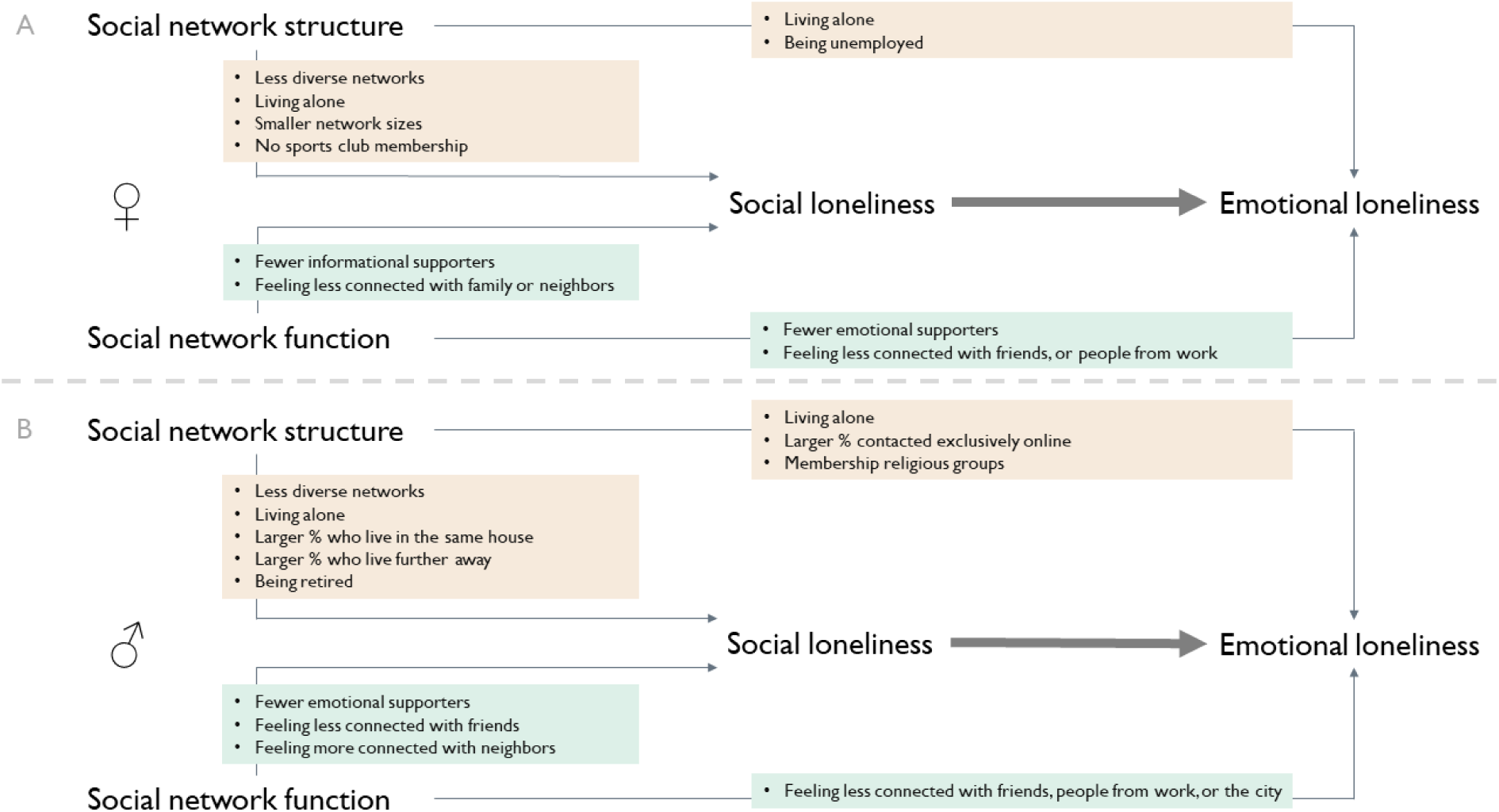
Overview of associations between social network aspects and moderate/severe social and emotional loneliness among men and women in the SaNAE study. Panel A shows associations for women and panel B shows association for men.

Sensitivity analyses showed similar results for the observed associated network aspects, when using another a cut-off value (≥1) for both social and emotional loneliness (supplementary table 2 and 3).

## 4. Discussion

This detailed evaluation of loneliness in Dutch adults aged 40 years and older, uniquely evaluated structure and function of social networks and did so in relation to social loneliness as well as emotional loneliness. Of men, 20% and 25% were moderate or severe socially and emotionally lonely, respectively. Of women, 16% and 32% were moderate or severe socially and emotionally lonely, respectively. More than half of the people who were socially lonely, were also emotionally lonely. Also a sizeable proportion (19%-24%) were ‘only’ moderate or severe emotionally lonely. By evaluating a wide range of structural and functional social network aspects, jointly, this study was able to identify the most important social network aspects for men and women, and for the two types of loneliness. These insights can be further used in the assessment of relevant social network aspects and strategies to prevent or alleviate loneliness.

Loneliness is not merely defined by either a structural or functional aspect and is certainly more than ‘just’ the lack of number of relationships or lack of social support. Striking differences were observed for social or emotional loneliness in this respect as well. Men and women in the current study were more moderately/severely socially lonely when their social network was less diverse (e.g., family-centered), in line with previous studies (Kemperman et al., 2019; Moorer & Suurmeijer, 2001; Wenger, 1997; Wenger & Tucker, 2002) or less dense (friends and family clustered less). Also, a small network was associated with social loneliness (women only) in line with previous studies (Dahlberg et al., 2018). Further, men and women were more likely to be socially lonely when they lived alone. Living alone is a well-known factor in loneliness and other adverse health outcomes (Brittain et al., 2017; Bu et al., 2020; Newall et al., 2009; Taube et al., 2013). Also important in social loneliness were feeling less connected to friends, fewer emotional supporters (for men only), fewer informational supporters (for women only), and not had a club membership.

Men and women were moderately/severely lonely when they also felt socially lonely, when they lived alone, when a larger proportion of their network members was contacted exclusively online (for men only) or when having fewer emotional supporters (for women only). Various metrics have been shown to be important in previous studies (Dahlberg et al., 2022), while this study thus also revealed novel insights.

Important to highlight for example is that less diverse and less dense social networks likely posed a risk for social or emotional loneliness in men and women, regardless the number of social relationships a person has. This has implications for assessing social network structure in relation to health, indicating the importance of taking network diversity and density into account. Also notable was that women who had fewer informational supporters and men who had fewer emotional supporters were more likely to be socially lonely, indicating that men and women may have different needs for social support from their social network. Finally, online contact (for men) was found detrimental for emotional loneliness, highlighting the value for in-person contact which was also observed in a qualitative study. (Steijvers et al., 2023)

Physical contact with social network members was reduced and was (temporarily) replaced with online contact during the COVID-19 pandemic. (Steijvers et al., 2022) Especially those who live alone were affected by these measures since online contact among individuals who live alone was in previous studies with negative emotions and loneliness. (Currin et al., 2022; Fingerman et al., 2021) Online contact might not be a full substitute for in person contact (Hawkley & Kocherginsky, 2018) and it has been demonstrated to decrease the level of feeling connected to others, which in turn negatively impacts health. (Holaday et al., 2022)

Social participation was also linked to social loneliness, as women in the current study who did not have a sports club membership more often were lonely. Previous studies have reported that women who exercise with other may do so for motivation and social conviviality (Allender et al., 2006), and exercising together stimulates to exercise more frequently. (Faulkner et al., 2021; Firestone et al., 2015; Ojiambo, 2013) Men in this study who were members of religious groups were more likely to be emotional lonely, and reasons are unknown and topic of further study.

### 4.1. Implications

To curb trends in loneliness is a key public health priority. This means to better identify relevant social network aspects in loneliness, and to better use these insights for designing strategies to prevent or alleviate loneliness. The current study provides insight into possible targets for the identification of loneliness and strategies to prevent or to alleviate loneliness. These results stress the importance to look beyond the number of social relationships and consider the rich variety of social network aspects and thereby take into account the sex differences.

Examples for better identification include a small network size, but also less diverse relationships. Employment status and living alone could function as a possible indicator of loneliness. Targeting social loneliness may also in part target emotional loneliness, but not completely as different social network aspects are important. In line with the work of Holt-Lunstad, we suggest that future research should focus on the development of social connection guidelines including the social network aspects identified in the current study. (Holt-Lunstad, 2023)

Examples for preventive strategies focused on network structure could be to help people to expand their network by adding new network members to also create a more diverse social network, and by strengthening connections between social network members to increase density. Club memberships (e.g., sport or music organizations) should be promoted and facilitated to enable people to meet new people in leisure activities. Strategies should further build on strengthening social network function for social support in existing social network members and relevant support in new members.

### 4.2. Strengths and limitations

A strength of the current study is that we used a large cohort study that uniquely and jointly assessed various structural and functional social network aspects in men and women, in combination with health-outcomes such as those evaluated here. To generate the social network aspects, a name generator questionnaire with name interpreter items was used, which is a useful method for measuring social networks in online surveys extracting large and diverse networks (Campbell & Lee, 1991). Furthermore, we evaluated the social network aspects separately for social and emotional loneliness and thereby identified which aspects are most important for each type of loneliness. Some limitations should be mentioned. Different answer categories for the De Jong Gierveld scale were used. Instead of three answer categories: ‘yes’, ‘more or less’, and ‘no’, a five-Likert scale was used ‘totally agree – totally disagree’. Reliability tests for social and emotional loneliness items were performed. Items were reliable (social loneliness items: α=0.911, and emotional loneliness items: α=0.794). Furthermore, due to the cross-sectional design of the current study, no conclusions can be drawn on the causality of the effects.

### 4.3. Conclusion

Our current study assessed structural and functional social network aspects associated with moderate/severe social and emotional loneliness for men and women separately and established that diverse and dense social networks, and emotional and informational social support are key factors associated with social and emotional loneliness. Preventive strategies to alleviate or prevent loneliness should focus on both structural and functional network aspects need to look beyond the number of social relationships and promote diverse and supporting social relationships.

## Data Availability

Data produced in the present study are available upon reasonable request to the authors

## 6. Supplementary files

**Supplementary table 1.**
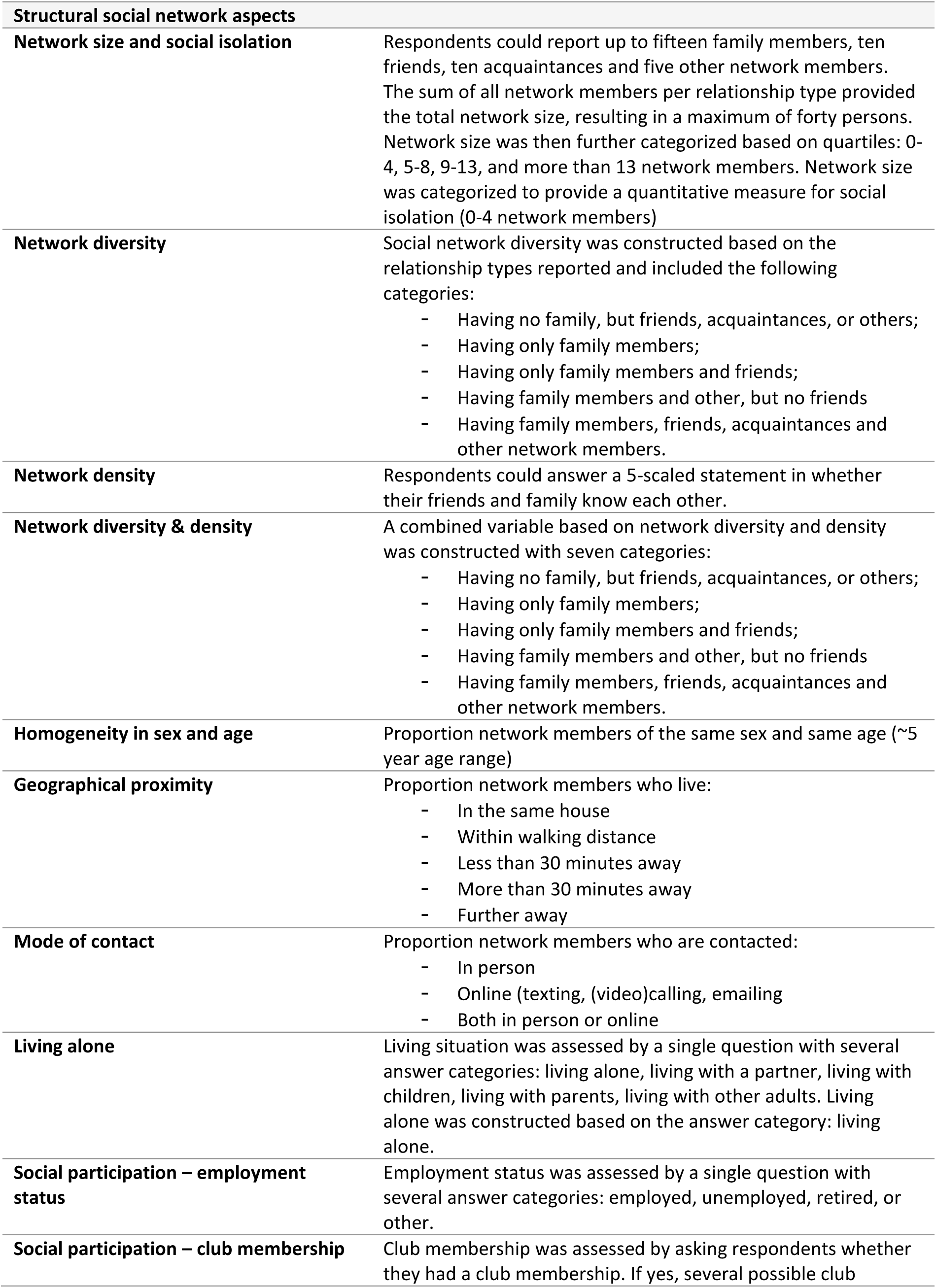
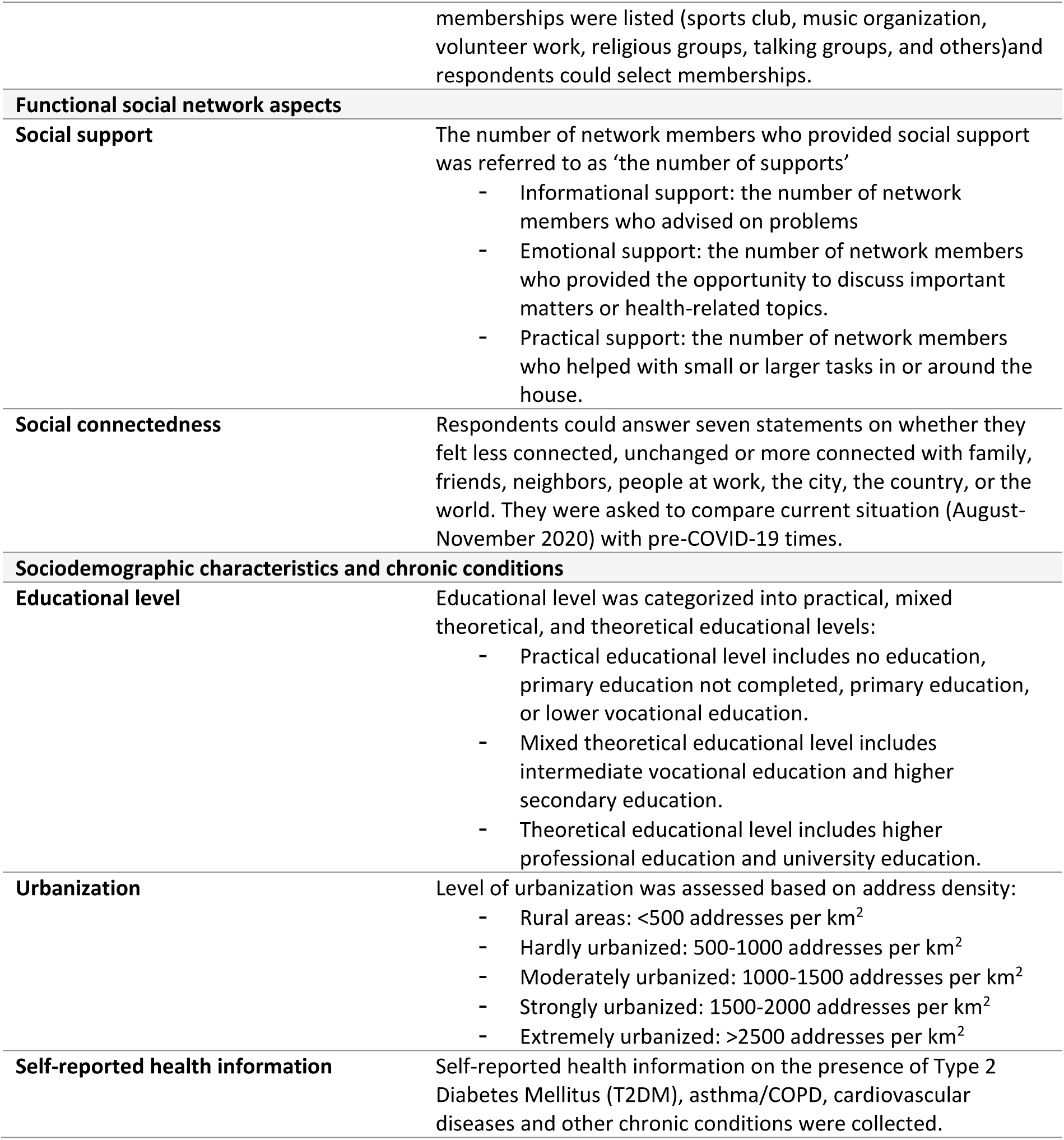
Overview of social network variables, sociodemographic variables, and chronic conditions.

**Supplementary table 2.**
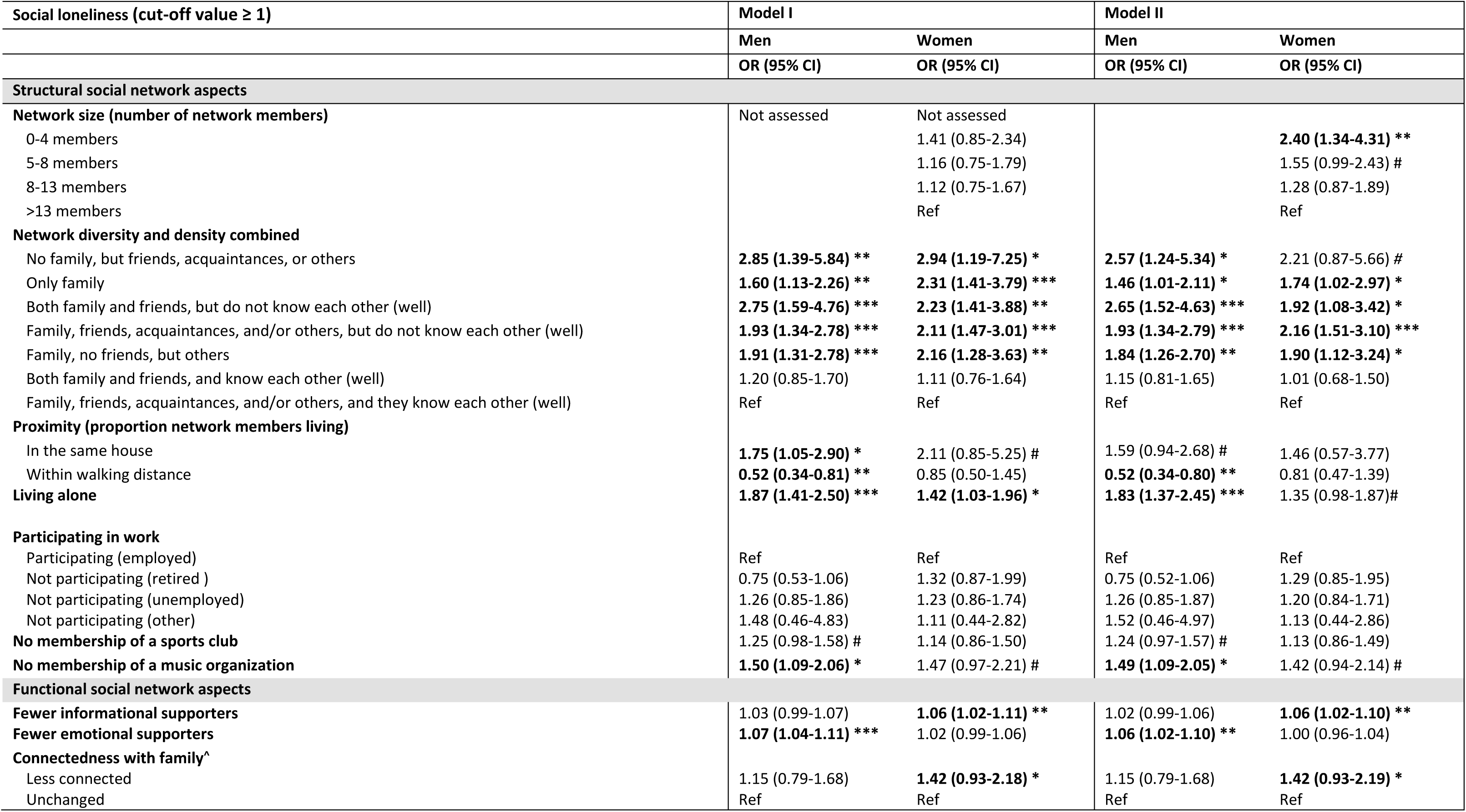
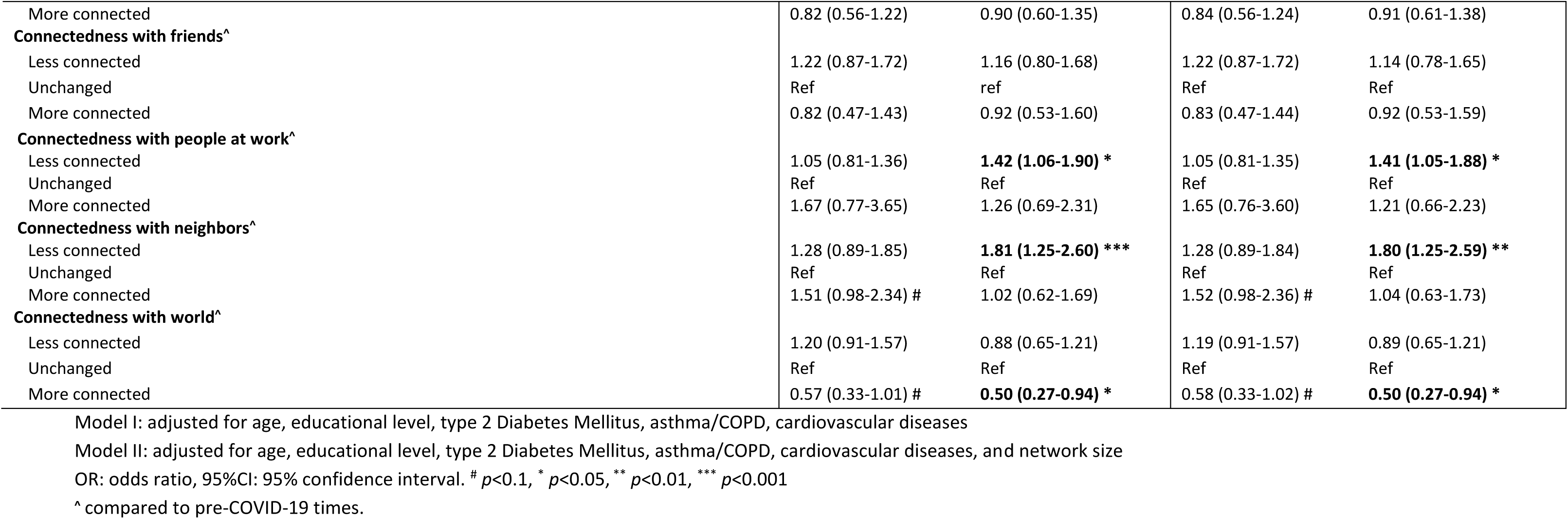
Multivariable logistic regression analyses between structural and functional social network aspects and social loneliness (cut-off value ≥ 1)

**Supplementary table 3.**
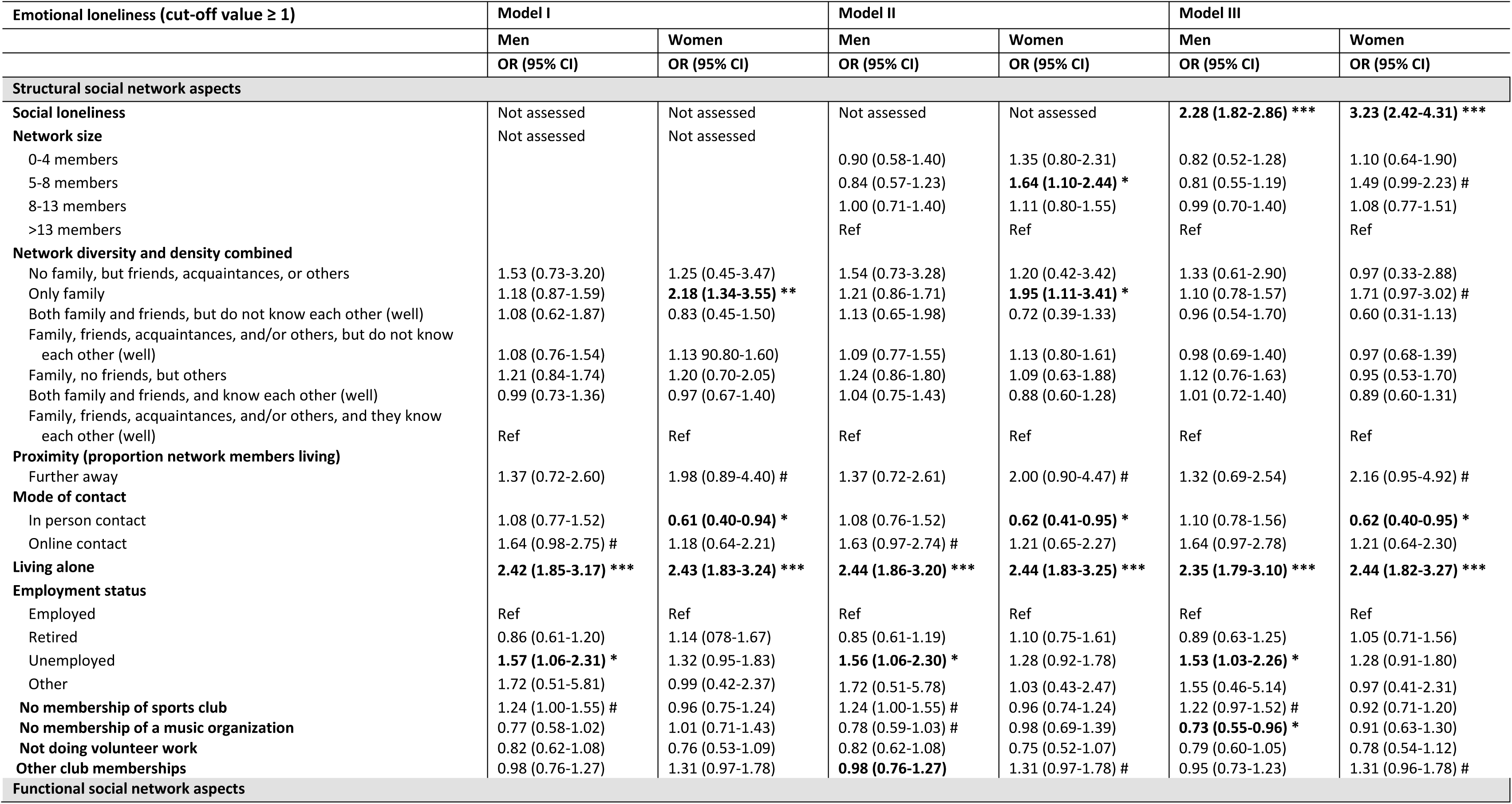
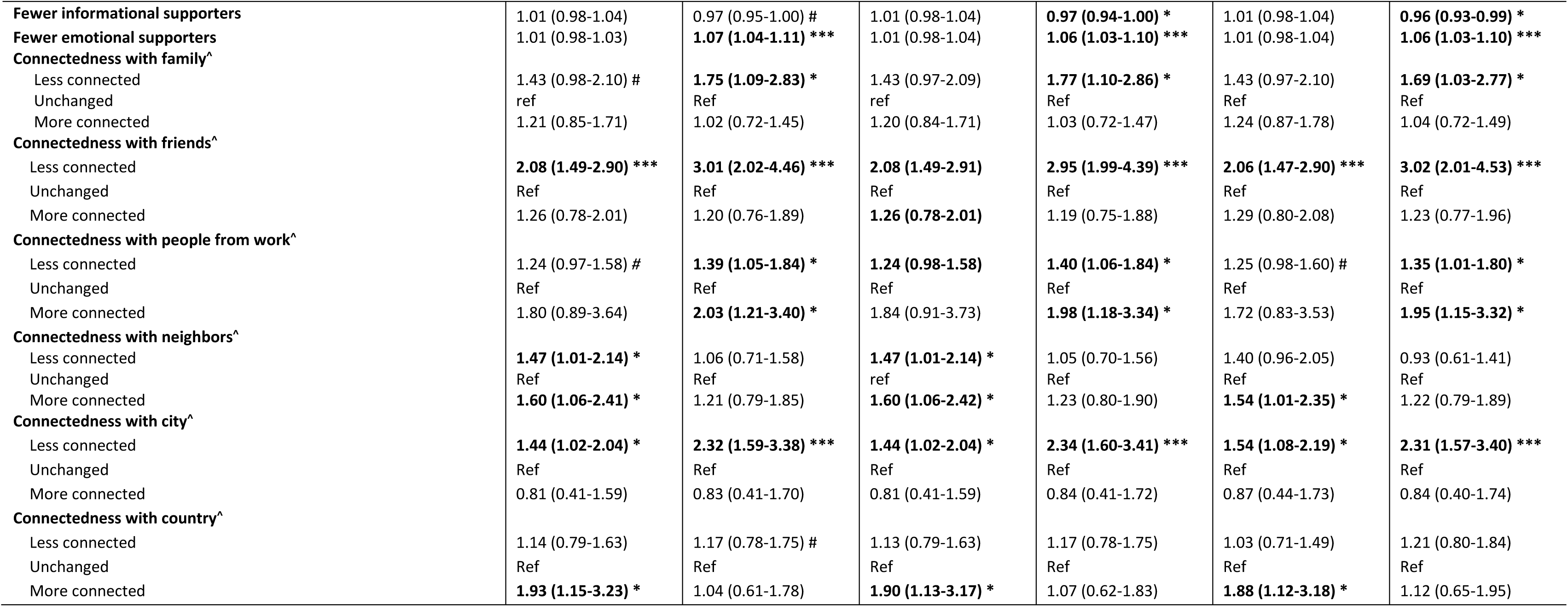
Multivariable logistic regression analyses between structural and functional social network aspects and emotional loneliness (cut-off value ≥ 1)

