## Supplementary Tables for "The role of social network structure and function in moderate and severe social and emotional loneliness: the Dutch SaNAE study in older adults"

### Supplementary files

**Supplementary table 1. Overview of social network variables, sociodemographic variables, and chronic conditions**

| Structural social network aspects | |
|---|---|
| **Network size and social isolation** | Respondents could report up to fifteen family members, ten friends, ten acquaintances and five other network members. The sum of all network members per relationship type provided the total network size, resulting in a maximum of forty persons. Network size was then further categorized based on quartiles: 0-4, 5-8, 9-13, and more than 13 network members. Network size was categorized to provide a quantitative measure for social isolation (0-4 network members) |
| **Network diversity** | Social network diversity was constructed based on the relationship types reported and included the following categories:<br>- Having no family, but friends, acquaintances, or others;<br>- Having only family members;<br>- Having only family members and friends;<br>- Having family members and other, but no friends<br>- Having family members, friends, acquaintances and other network members. |
| **Network density** | Respondents could answer a 5-scaled statement in whether their friends and family know each other. |
| **Network diversity & density** | A combined variable based on network diversity and density was constructed with seven categories:<br>- Having no family, but friends, acquaintances, or others;<br>- Having only family members;<br>- Having only family members and friends;<br>- Having family members and other, but no friends<br>- Having family members, friends, acquaintances and other network members. |
| **Homogeneity in sex and age** | Proportion network members of the same sex and same age (~5 year age range) |
| **Geographical proximity** | Proportion network members who live:<br>- In the same house<br>- Within walking distance<br>- Less than 30 minutes away<br>- More than 30 minutes away<br>- Further away |
| **Mode of contact** | Proportion network members who are contacted:<br>- In person<br>- Online (texting, (video)calling, emailing<br>- Both in person or online |
| **Living alone** | Living situation was assessed by a single question with several answer categories: living alone, living with a partner, living with children, living with parents, living with other adults. Living alone was constructed based on the answer category: living alone. |
| **Social participation – employment status** | Employment status was assessed by a single question with several answer categories: employed, unemployed, retired, or other. |
| **Social participation – club membership** | Club membership was assessed by asking respondents whether they had a club membership. If yes, several possible club |

| | |
|---|---|
| | memberships were listed (sports club, music organization, volunteer work, religious groups, talking groups, and others)and respondents could select memberships. |
| **Functional social network aspects** | |
| **Social support** | The number of network members who provided social support was referred to as ‘the number of supports’<br>- Informational support: the number of network members who advised on problems<br>- Emotional support: the number of network members who provided the opportunity to discuss important matters or health-related topics.<br>- Practical support: the number of network members who helped with small or larger tasks in or around the house. |
| **Social connectedness** | Respondents could answer seven statements on whether they felt less connected, unchanged or more connected with family, friends, neighbors, people at work, the city, the country, or the world. They were asked to compare current situation (August-November 2020) with pre-COVID-19 times. |
| **Sociodemographic characteristics and chronic conditions** | |
| **Educational level** | Educational level was categorized into practical, mixed theoretical, and theoretical educational levels:<br>- Practical educational level includes no education, primary education not completed, primary education, or lower vocational education.<br>- Mixed theoretical educational level includes intermediate vocational education and higher secondary education.<br>- Theoretical educational level includes higher professional education and university education. |
| **Urbanization** | Level of urbanization was assessed based on address density:<br>- Rural areas: <500 addresses per $km^2$<br>- Hardly urbanized: 500-1000 addresses per $km^2$<br>- Moderately urbanized: 1000-1500 addresses per $km^2$<br>- Strongly urbanized: 1500-2000 addresses per $km^2$<br>- Extremely urbanized: >2500 addresses per $km^2$ |
| **Self-reported health information** | Self-reported health information on the presence of Type 2 Diabetes Mellitus (T2DM), asthma/COPD, cardiovascular diseases and other chronic conditions were collected. |

**Supplementary table 2. Multivariable logistic regression analyses between structural and functional social network aspects and social loneliness (cut-off value ≥ 1)**

| Social loneliness (cut-off value ≥ 1) | Model I | | Model II | |
|---|---|---|---|---|
| | **Men** | **Women** | **Men** | **Women** |
| | **OR (95% CI)** | **OR (95% CI)** | **OR (95% CI)** | **OR (95% CI)** |
| **Structural social network aspects** | | | | |
| **Network size (number of network members)** | Not assessed | Not assessed | | |
| 0-4 members | | 1.41 (0.85-2.34) | | **2.40 (1.34-4.31)** ** |
| 5-8 members | | 1.16 (0.75-1.79) | | 1.55 (0.99-2.43) # |
| 8-13 members | | 1.12 (0.75-1.67) | | 1.28 (0.87-1.89) |
| >13 members | | Ref | | Ref |
| **Network diversity and density combined** | | | | |
| No family, but friends, acquaintances, or others | **2.85 (1.39-5.84)** ** | **2.94 (1.19-7.25)** * | **2.57 (1.24-5.34)** * | 2.21 (0.87-5.66) # |
| Only family | **1.60 (1.13-2.26)** ** | **2.31 (1.41-3.79)** *** | **1.46 (1.01-2.11)** * | **1.74 (1.02-2.97)** * |
| Both family and friends, but do not know each other (well) | **2.75 (1.59-4.76)** *** | **2.23 (1.41-3.88)** ** | **2.65 (1.52-4.63)** *** | **1.92 (1.08-3.42)** * |
| Family, friends, acquaintances, and/or others, but do not know each other (well) | **1.93 (1.34-2.78)** *** | **2.11 (1.47-3.01)** *** | **1.93 (1.34-2.79)** *** | **2.16 (1.51-3.10)** *** |
| Family, no friends, but others | **1.91 (1.31-2.78)** *** | **2.16 (1.28-3.63)** ** | **1.84 (1.26-2.70)** ** | **1.90 (1.12-3.24)** * |
| Both family and friends, and know each other (well) | 1.20 (0.85-1.70) | 1.11 (0.76-1.64) | 1.15 (0.81-1.65) | 1.01 (0.68-1.50) |
| Family, friends, acquaintances, and/or others, and they know each other (well) | Ref | Ref | Ref | Ref |
| **Proximity (proportion network members living)** | | | | |
| In the same house | **1.75 (1.05-2.90)** * | 2.11 (0.85-5.25) # | 1.59 (0.94-2.68) # | 1.46 (0.57-3.77) |
| Within walking distance | **0.52 (0.34-0.81)** ** | 0.85 (0.50-1.45) | **0.52 (0.34-0.80)** ** | 0.81 (0.47-1.39) |
| **Living alone** | **1.87 (1.41-2.50)** *** | **1.42 (1.03-1.96)** * | **1.83 (1.37-2.45)** *** | 1.35 (0.98-1.87)# |
| **Participating in work** | | | | |
| Participating (employed) | Ref | Ref | Ref | Ref |
| Not participating (retired ) | 0.75 (0.53-1.06) | 1.32 (0.87-1.99) | 0.75 (0.52-1.06) | 1.29 (0.85-1.95) |
| Not participating (unemployed) | 1.26 (0.85-1.86) | 1.23 (0.86-1.74) | 1.26 (0.85-1.87) | 1.20 (0.84-1.71) |
| Not participating (other) | 1.48 (0.46-4.83) | 1.11 (0.44-2.82) | 1.52 (0.46-4.97) | 1.13 (0.44-2.86) |
| **No membership of a sports club** | 1.25 (0.98-1.58) # | 1.14 (0.86-1.50) | 1.24 (0.97-1.57) # | 1.13 (0.86-1.49) |
| **No membership of a music organization** | **1.50 (1.09-2.06)** * | 1.47 (0.97-2.21) # | **1.49 (1.09-2.05)** * | 1.42 (0.94-2.14) # |
| **Functional social network aspects** | | | | |
| **Fewer informational supporters** | 1.03 (0.99-1.07) | **1.06 (1.02-1.11)** ** | 1.02 (0.99-1.06) | **1.06 (1.02-1.10)** ** |
| **Fewer emotional supporters** | **1.07 (1.04-1.11)** *** | 1.02 (0.99-1.06) | **1.06 (1.02-1.10)** ** | 1.00 (0.96-1.04) |
| **Connectedness with family^** | | | | |
| Less connected | 1.15 (0.79-1.68) | **1.42 (0.93-2.18)** * | 1.15 (0.79-1.68) | **1.42 (0.93-2.19)** * |
| Unchanged | Ref | Ref | Ref | Ref |

| | | | | |
|---|---|---|---|---|
| More connected | 0.82 (0.56-1.22) | 0.90 (0.60-1.35) | 0.84 (0.56-1.24) | 0.91 (0.61-1.38) |
| **Connectedness with friends**^ | | | | |
| Less connected | 1.22 (0.87-1.72) | 1.16 (0.80-1.68) | 1.22 (0.87-1.72) | 1.14 (0.78-1.65) |
| Unchanged | Ref | ref | Ref | Ref |
| More connected | 0.82 (0.47-1.43) | 0.92 (0.53-1.60) | 0.83 (0.47-1.44) | 0.92 (0.53-1.59) |
| **Connectedness with people at work**^ | | | | |
| Less connected | 1.05 (0.81-1.36) | **1.42 (1.06-1.90) *** | 1.05 (0.81-1.35) | **1.41 (1.05-1.88) *** |
| Unchanged | Ref | Ref | Ref | Ref |
| More connected | 1.67 (0.77-3.65) | 1.26 (0.69-2.31) | 1.65 (0.76-3.60) | 1.21 (0.66-2.23) |
| **Connectedness with neighbors**^ | | | | |
| Less connected | 1.28 (0.89-1.85) | **1.81 (1.25-2.60) ***** | 1.28 (0.89-1.84) | **1.80 (1.25-2.59) **** |
| Unchanged | Ref | Ref | Ref | Ref |
| More connected | 1.51 (0.98-2.34) # | 1.02 (0.62-1.69) | 1.52 (0.98-2.36) # | 1.04 (0.63-1.73) |
| **Connectedness with world**^ | | | | |
| Less connected | 1.20 (0.91-1.57) | 0.88 (0.65-1.21) | 1.19 (0.91-1.57) | 0.89 (0.65-1.21) |
| Unchanged | Ref | Ref | Ref | Ref |
| More connected | 0.57 (0.33-1.01) # | **0.50 (0.27-0.94) *** | 0.58 (0.33-1.02) # | **0.50 (0.27-0.94) *** |

Model I: adjusted for age, educational level, type 2 Diabetes Mellitus, asthma/COPD, cardiovascular diseases

Model II: adjusted for age, educational level, type 2 Diabetes Mellitus, asthma/COPD, cardiovascular diseases, and network size

OR: odds ratio, 95%CI: 95% confidence interval. $^{\#}$ $p$<0.1, $^{*}$ $p$<0.05, $^{**}$ $p$<0.01, $^{***}$ $p$<0.001

^ compared to pre-COVID-19 times.

**Supplementary table 3. Multivariable logistic regression analyses between structural and functional social network aspects and emotional loneliness (cut-off value ≥ 1)**

| Emotional loneliness (cut-off value ≥ 1) | Model I | | Model II | | Model III | |
|---|---|---|---|---|---|---|
| | Men | Women | Men | Women | Men | Women |
| | OR (95% CI) | OR (95% CI) | OR (95% CI) | OR (95% CI) | OR (95% CI) | OR (95% CI) |
| **Structural social network aspects** | | | | | | |
| **Social loneliness** | Not assessed | Not assessed | Not assessed | Not assessed | **2.28 (1.82-2.86) ***** | **3.23 (2.42-4.31) ***** |
| **Network size** | Not assessed | Not assessed | | | | |
| 0-4 members | | | 0.90 (0.58-1.40) | 1.35 (0.80-2.31) | 0.82 (0.52-1.28) | 1.10 (0.64-1.90) |
| 5-8 members | | | 0.84 (0.57-1.23) | **1.64 (1.10-2.44) *** | 0.81 (0.55-1.19) | 1.49 (0.99-2.23) # |
| 8-13 members | | | 1.00 (0.71-1.40) | 1.11 (0.80-1.55) | 0.99 (0.70-1.40) | 1.08 (0.77-1.51) |
| >13 members | | | Ref | Ref | Ref | Ref |
| **Network diversity and density combined** | | | | | | |
| No family, but friends, acquaintances, or others | 1.53 (0.73-3.20) | 1.25 (0.45-3.47) | 1.54 (0.73-3.28) | 1.20 (0.42-3.42) | 1.33 (0.61-2.90) | 0.97 (0.33-2.88) |
| Only family | 1.18 (0.87-1.59) | **2.18 (1.34-3.55) **** | 1.21 (0.86-1.71) | **1.95 (1.11-3.41) *** | 1.10 (0.78-1.57) | 1.71 (0.97-3.02) # |
| Both family and friends, but do not know each other (well) | 1.08 (0.62-1.87) | 0.83 (0.45-1.50) | 1.13 (0.65-1.98) | 0.72 (0.39-1.33) | 0.96 (0.54-1.70) | 0.60 (0.31-1.13) |
| Family, friends, acquaintances, and/or others, but do not know each other (well) | 1.08 (0.76-1.54) | 1.13 90.80-1.60) | 1.09 (0.77-1.55) | 1.13 (0.80-1.61) | 0.98 (0.69-1.40) | 0.97 (0.68-1.39) |
| Family, no friends, but others | 1.21 (0.84-1.74) | 1.20 (0.70-2.05) | 1.24 (0.86-1.80) | 1.09 (0.63-1.88) | 1.12 (0.76-1.63) | 0.95 (0.53-1.70) |
| Both family and friends, and know each other (well) | 0.99 (0.73-1.36) | 0.97 (0.67-1.40) | 1.04 (0.75-1.43) | 0.88 (0.60-1.28) | 1.01 (0.72-1.40) | 0.89 (0.60-1.31) |
| Family, friends, acquaintances, and/or others, and they know each other (well) | Ref | Ref | Ref | Ref | Ref | Ref |
| **Proximity (proportion network members living)** | | | | | | |
| Further away | 1.37 (0.72-2.60) | 1.98 (0.89-4.40) # | 1.37 (0.72-2.61) | 2.00 (0.90-4.47) # | 1.32 (0.69-2.54) | 2.16 (0.95-4.92) # |
| **Mode of contact** | | | | | | |
| In person contact | 1.08 (0.77-1.52) | **0.61 (0.40-0.94) *** | 1.08 (0.76-1.52) | **0.62 (0.41-0.95) *** | 1.10 (0.78-1.56) | **0.62 (0.40-0.95) *** |
| Online contact | 1.64 (0.98-2.75) # | 1.18 (0.64-2.21) | 1.63 (0.97-2.74) # | 1.21 (0.65-2.27) | 1.64 (0.97-2.78) | 1.21 (0.64-2.30) |
| **Living alone** | **2.42 (1.85-3.17) ***** | **2.43 (1.83-3.24) ***** | **2.44 (1.86-3.20) ***** | **2.44 (1.83-3.25) ***** | **2.35 (1.79-3.10) ***** | **2.44 (1.82-3.27) ***** |
| **Employment status** | | | | | | |
| Employed | Ref | Ref | Ref | Ref | Ref | Ref |
| Retired | 0.86 (0.61-1.20) | 1.14 (078-1.67) | 0.85 (0.61-1.19) | 1.10 (0.75-1.61) | 0.89 (0.63-1.25) | 1.05 (0.71-1.56) |
| Unemployed | **1.57 (1.06-2.31) *** | 1.32 (0.95-1.83) | **1.56 (1.06-2.30) *** | 1.28 (0.92-1.78) | **1.53 (1.03-2.26) *** | 1.28 (0.91-1.80) |
| Other | 1.72 (0.51-5.81) | 0.99 (0.42-2.37) | 1.72 (0.51-5.78) | 1.03 (0.43-2.47) | 1.55 (0.46-5.14) | 0.97 (0.41-2.31) |
| **No membership of sports club** | 1.24 (1.00-1.55) # | 0.96 (0.75-1.24) | 1.24 (1.00-1.55) # | 0.96 (0.74-1.24) | 1.22 (0.97-1.52) # | 0.92 (0.71-1.20) |
| **No membership of a music organization** | 0.77 (0.58-1.02) | 1.01 (0.71-1.43) | 0.78 (0.59-1.03) # | 0.98 (0.69-1.39) | **0.73 (0.55-0.96) *** | 0.91 (0.63-1.30) |
| **Not doing volunteer work** | 0.82 (0.62-1.08) | 0.76 (0.53-1.09) | 0.82 (0.62-1.08) | 0.75 (0.52-1.07) | 0.79 (0.60-1.05) | 0.78 (0.54-1.12) |
| **Other club memberships** | 0.98 (0.76-1.27) | 1.31 (0.97-1.78) | **0.98 (0.76-1.27)** | 1.31 (0.97-1.78) # | 0.95 (0.73-1.23) | 1.31 (0.96-1.78) # |
| **Functional social network aspects** | | | | | | |

| | | | | | | |
|---|---|---|---|---|---|---|
| **Fewer informational supporters** | 1.01 (0.98-1.04) | 0.97 (0.95-1.00) # | 1.01 (0.98-1.04) | **0.97 (0.94-1.00) *** | 1.01 (0.98-1.04) | **0.96 (0.93-0.99) *** |
| **Fewer emotional supporters** | 1.01 (0.98-1.03) | **1.07 (1.04-1.11) ***** | 1.01 (0.98-1.04) | **1.06 (1.03-1.10) ***** | 1.01 (0.98-1.04) | **1.06 (1.03-1.10) ***** |
| **Connectedness with family^** | | | | | | |
| Less connected | 1.43 (0.98-2.10) # | **1.75 (1.09-2.83) *** | 1.43 (0.97-2.09) | **1.77 (1.10-2.86) *** | 1.43 (0.97-2.10) | **1.69 (1.03-2.77) *** |
| Unchanged | ref | Ref | ref | Ref | Ref | Ref |
| More connected | 1.21 (0.85-1.71) | 1.02 (0.72-1.45) | 1.20 (0.84-1.71) | 1.03 (0.72-1.47) | 1.24 (0.87-1.78) | 1.04 (0.72-1.49) |
| **Connectedness with friends^** | | | | | | |
| Less connected | **2.08 (1.49-2.90) ***** | **3.01 (2.02-4.46) ***** | **2.08 (1.49-2.91)** | **2.95 (1.99-4.39) ***** | **2.06 (1.47-2.90) ***** | **3.02 (2.01-4.53) ***** |
| Unchanged | Ref | Ref | Ref | Ref | Ref | Ref |
| More connected | 1.26 (0.78-2.01) | 1.20 (0.76-1.89) | **1.26 (0.78-2.01)** | 1.19 (0.75-1.88) | 1.29 (0.80-2.08) | 1.23 (0.77-1.96) |
| **Connectedness with people from work^** | | | | | | |
| Less connected | 1.24 (0.97-1.58) # | **1.39 (1.05-1.84) *** | **1.24 (0.98-1.58)** | **1.40 (1.06-1.84) *** | 1.25 (0.98-1.60) # | **1.35 (1.01-1.80) *** |
| Unchanged | Ref | Ref | Ref | Ref | Ref | Ref |
| More connected | 1.80 (0.89-3.64) | **2.03 (1.21-3.40) *** | 1.84 (0.91-3.73) | **1.98 (1.18-3.34) *** | 1.72 (0.83-3.53) | **1.95 (1.15-3.32) *** |
| **Connectedness with neighbors^** | | | | | | |
| Less connected | **1.47 (1.01-2.14) *** | 1.06 (0.71-1.58) | **1.47 (1.01-2.14) *** | 1.05 (0.70-1.56) | 1.40 (0.96-2.05) | 0.93 (0.61-1.41) |
| Unchanged | Ref | Ref | ref | Ref | Ref | Ref |
| More connected | **1.60 (1.06-2.41) *** | 1.21 (0.79-1.85) | **1.60 (1.06-2.42) *** | 1.23 (0.80-1.90) | **1.54 (1.01-2.35) *** | 1.22 (0.79-1.89) |
| **Connectedness with city^** | | | | | | |
| Less connected | **1.44 (1.02-2.04) *** | **2.32 (1.59-3.38) ***** | **1.44 (1.02-2.04) *** | **2.34 (1.60-3.41) ***** | **1.54 (1.08-2.19) *** | **2.31 (1.57-3.40) ***** |
| Unchanged | Ref | Ref | Ref | Ref | Ref | Ref |
| More connected | 0.81 (0.41-1.59) | 0.83 (0.41-1.70) | 0.81 (0.41-1.59) | 0.84 (0.41-1.72) | 0.87 (0.44-1.73) | 0.84 (0.40-1.74) |
| **Connectedness with country^** | | | | | | |
| Less connected | 1.14 (0.79-1.63) | 1.17 (0.78-1.75) # | 1.13 (0.79-1.63) | 1.17 (0.78-1.75) | 1.03 (0.71-1.49) | 1.21 (0.80-1.84) |
| Unchanged | Ref | Ref | Ref | Ref | Ref | Ref |
| More connected | **1.93 (1.15-3.23) *** | 1.04 (0.61-1.78) | **1.90 (1.13-3.17) *** | 1.07 (0.62-1.83) | **1.88 (1.12-3.18) *** | 1.12 (0.65-1.95) |
